# A national cross-sectional survey of public perceptions, knowledge, and behaviors during the COVID-19 pandemic

**DOI:** 10.1101/2020.07.07.20147413

**Authors:** Jeanna Parsons Leigh, Kirsten Fiest, Rebecca Brundin-Mather, Kara Plotnikoff, Andrea Soo, Emma E. Sypes, Liam Whalen-Browne, Sofia B. Ahmed, Karen E.A. Burns, Alison Fox-Robichaud, Shelly Kupsch, Shelly Longmore, Srinivas Murthy, Daniel J. Niven, Bram Rochwerg, Henry T. Stelfox

## Abstract

**Introduction:** Efforts to mitigate the global spread of the severe acute respiratory syndrome coronavirus 2 (SARS-CoV-2) have largely relied on broad compliance with public health recommendations yet navigating the high volume of evolving information and misinformation related to SARS-CoV-2 can be challenging. We assessed national public perceptions (e.g., severity, concerns, health), knowledge (e.g., transmission, information sources), and behaviors (e.g., physical distancing) related to COVID-19 in Canada to understand public perspectives and inform future public health initiatives.

**Methods:** We administered a national online survey with the goal of obtaining responses from 2000 adults residing in Canada. Respondent sampling was stratified by age, sex, and region. We used descriptive statistics to summarize respondent characteristics and tested for significant overall regional differences using chi-squared tests and t-tests, as appropriate.

**Results:** We collected 1,996 eligible questionnaires between April 26^th^ and May 1^st^, 2020. One-fifth (20%) of respondents knew someone diagnosed with COVID-19, but few had tested positive themselves (0.6%). Negative impacts of pandemic conditions were evidenced in several areas, including concerns about healthcare (e.g. sufficient equipment, 52%), pandemic stress (45%), and worsening social (49%) and mental/emotional (39%) health. Most respondents (88%) felt they had good to excellent knowledge of virus transmission, and predominantly accessed (74%) and trusted (60%) Canadian news television, newspapers/magazines, or non-government news websites for COVID-19 information. We found high compliance with distancing measures (80% either self-isolating or always physical distancing). We identified regional differences in perceptions, knowledge, and behaviors related to COVID-19.

**Discussion:** We found that knowledge about COVID-19 is largely acquired through domestic news sources, which may explain high self-reported compliance with prevention measures. The results highlight the broader impact of a pandemic on the general public’s overall health and wellbeing, outside of personal infection. The study findings should be used to inform public health communications during COVID-19 and future pandemics.

## Introduction

Since the emergence of the severe acute respiratory syndrome coronavirus 2 (SARS-CoV-2) in December 2019,(1) the global public has been inundated with information related to the rapidly evolving Coronavirus Disease 2019 (COVID-19) pandemic.(2) Organizations such as the World Health Organization (WHO),(3) worldwide public health networks,(4) and government public health agencies(5) have used multiple media platforms (e.g., internet, television, radio, print) in attempts to keep the public informed of emerging details and public health recommendations. In Canada, this messaging has included mitigation strategies such as appropriate hand and face hygiene practices,(5) physical distancing policies including closing non-essential business and public spaces,(5) restrictions and limitations on visitation in hospitals and long-term care facilities,(6) and travel restrictions.(5) Effective and transparent communication of evolving information related to COVID-19 is needed to ensure the public understands how and why to adapt their behaviors to bolster public safety.(7) However, the influx of COVID-19 information and widespread circulation and exchange of misinformation (i.e., false or inaccurate information)(8, 9) have been linked to increased public fear,(10) under-use of health services,(11) and distrust in government messaging(12)–a phenomenon the WHO has characterized as an ‘infodemic’ (i.e., when the proliferation of information about a problem detracts from possible solutions).(9)

Effective pandemic management is dependent on understanding public views and behaviors, including concerns, frequently used and trusted sources of information, and motivations to observe or violate public health mandates.(7, 13) Countries around the world have used online cross-sectional surveys to rapidly assess public awareness, understand health behaviors, and identify sources of information and misinformation during COVID-19.(13-17) A survey of the American and British public early in the pandemic indicated adequate public awareness of disease transmission but a lack of understanding of appropriate preventative measures as well as high uptake of common misconceptions which were circulating on social media.(18) Public surveys have also revealed associations between socio-cultural factors (e.g., gender, socioeconomic status, education) and COVID-19 related levels of concern and knowledge.(19, 20) Evidence collected from public surveys(14, 16-18, 20) may inform the development of targeted public health messaging and track uptake of new information;(21) however, to date no comprehensive surveys investigating public perspectives and behaviors related to COVID-19 in Canada have been published. We conducted a national survey of adults residing in Canada to gain a better understanding of public perceptions (e.g. greatest concerns, views on government response), knowledge (e.g. sources, accuracy), and behaviors (e.g. prevention measures) related to the COVID-19 pandemic to inform future public health messaging and initiatives.

## Methods

### Study Design

We developed a cross-sectional, online, anonymous survey and contracted Ipsos Incorporated (https://www.ipsos.com/en-ca), a world-wide market research and polling firm, to administer it across Canada.

### Survey Design

#### Questionnaire Design and Testing

We iteratively synthesized a comprehensive list of questions based on broad content areas reported in previously published survey research on pandemics(22-25) and in current COVID-19 public opinion polls.(15) We subsequently invited seven members of the research team (co-investigators, research assistants, and patient partners) to provide feedback on question format, comprehensiveness, clarity, and flow.(26) We refined the questionnaire based on feedback.

The questionnaire domains and sub-domains are illustrated in S1 Fig. Question types included 5-point unipolar scales (e.g., 1=not at all/poor, 5=extremely/excellent), 7-point bipolar agreement scales (1=strongly disagree to 7=strongly agree), single-response multiple choice, and multiple response multiple choice. We randomized the order of the response options to reduce response selection bias.(26) We compared respondents’ retrospective ratings of five domains of overall health (mental/emotional, physical, social, economic, spiritual) at the start of 2020 to ratings of their health status at the time of data collection, with differences categorized into ‘worse’, ‘same’, or ‘better’. We provided respondents with definitions for self-isolation and social/physical distancing. Self-isolation was defined as *“separating yourself from others, including those within your home, with the purpose of preventing the spread of the virus (whether diagnosed or undiagnosed, with or without symptoms”* and social/physical distancing defined as *“limiting your time in spaces occupied by others, including reducing trips to visit others in person and reducing time spent in public spaces.”*

To ascertain whether the questionnaire could be completed within 15-minutes, we piloted it with a sample of 104 Canadians. No changes to the questionnaire were made and we therefore included the pilot responses in the final data set. The questionnaire was optimized for ‘device agnosticism’ to ensure its compatibility across most systems (e.g., mobile phone, computer, tablet).

### Questionnaire Formatting

The final questionnaire (see S1 Appendix) was formatted in English and French and consisted of 21 demographic and 46 COVID-19-related questions covering three overarching domains—perceptions, knowledge, and behaviors—important in gauging public response to, and government management of, the COVID-19 pandemic.

### Questionnaire Administration

The questionnaire was distributed electronically through Ipsos’ proprietary iSay panel of approximately 250,000 Canadians using direct email and social media posts. We included eligible panelists that were adults (≥ 18 years), lived in Canada, and were able to read English or French. We screened respondents by age (18-34, 35-55, >55), sex (female/male), and provincially defined regions (British Columbia, Alberta, Saskatchewan/Manitoba, Ontario, Québec, and Maritimes) to ensure population representation based on 2016 census data.(27) We excluded abandoned or incomplete questionnaires. Respondents received Ipsos reward points after completing the questionnaire; points are accumulated and redeemed for gift cards and merchandise.

### Sample Size Calculations

We derived a minimum sample size estimate of 385 based on a normal approximation to the binomial distribution with a finite population correction applied(28) (assuming an observed proportion of respondents selecting a specific response option of 50%) that incorporated population size (∼36.3 million in Canada), a 95% confidence level and a margin of error of 5%. We elected to collect 2,000 questionnaires to allow for subgroup analyses and calculated the associated margin of error to be +/-2.2% at a 95% confidence level.

### Data Analysis

We used descriptive statistics (frequencies (percent) or means (standard deviation)) to summarize respondent characteristics. We weighted responses by age, sex, and regional population estimates derived from 2016 census data.(27) Likert scales were reported as frequencies with percent for each point on the scale. We collapsed seven-point scale questions were collapsed into 5-point scales (strongly agree collapsed with agree, and strongly disagree collapsed with disagree) for analysis. We tested for differences between regions using chi-squared tests. We conducted all quantitative data analyses using SPSS, version 23 and R, version 3.5.1.(29) We used the R package “survey”(30) was used to obtain weighted descriptive statistics and chi-squared tests, version 3.36. Statistical significance was set at α=0.05.

### Ethical Oversight

Dalhousie University (#2020-5121) and the University of Calgary (#20-0538) Research Ethics Boards approved this study. Prior to entering the questionnaire, respondents reviewed an informed consent page; consent was implied by completing the questionnaire.

## Results

We collected data from April 26^th^ to May 1^st^, 2020. We excluded four respondents who reported being unaware of the current COVID-19 pandemic, resulting in a final sample of 1,996 respondents. On the last date data was collected (May 1^st^) there were 56,158 confirmed cases of COVID-19 in Canada; 83% of the cases were in the two most populated provinces, Québec (51%) and Ontario (32%).

### Respondent Characteristics

Only 12 (0.6%, 95% Confidence Interval (CI) 0.4%-1.0%) respondents reported ever testing positive for COVID-19. Most (n=1,855, 93.1%, 95%CI 91.9%-94.0%) were either uncertain or believed they had not contracted COVID-19; one-fifth (n=410, 20.6%) reported personally knowing someone diagnosed with COVID-19. Respondents were on average 49.4 (95%CI 48.6-50.2) years old. Just over half (n=1,018, 51.0%, 95%CI 48.8%-53.2%) were women, and 61 percent of all respondents were currently partnered (n=1216, 95%CI 59.2%-63.5%). Three-quarters of respondents reported a household income under 100,000 CND (n=1258, 71.8%, 95%CI 69.6%-73.9%). Just over one-half (n=563, 50.1%) of the 1001 employed respondents were working in a job deemed essential and 14 percent (n=143) of unemployed respondents (n=995) reported their unemployment being a direct result of COVID-19. Respondent characteristics are summarized in Table 1.

**Table 1.**
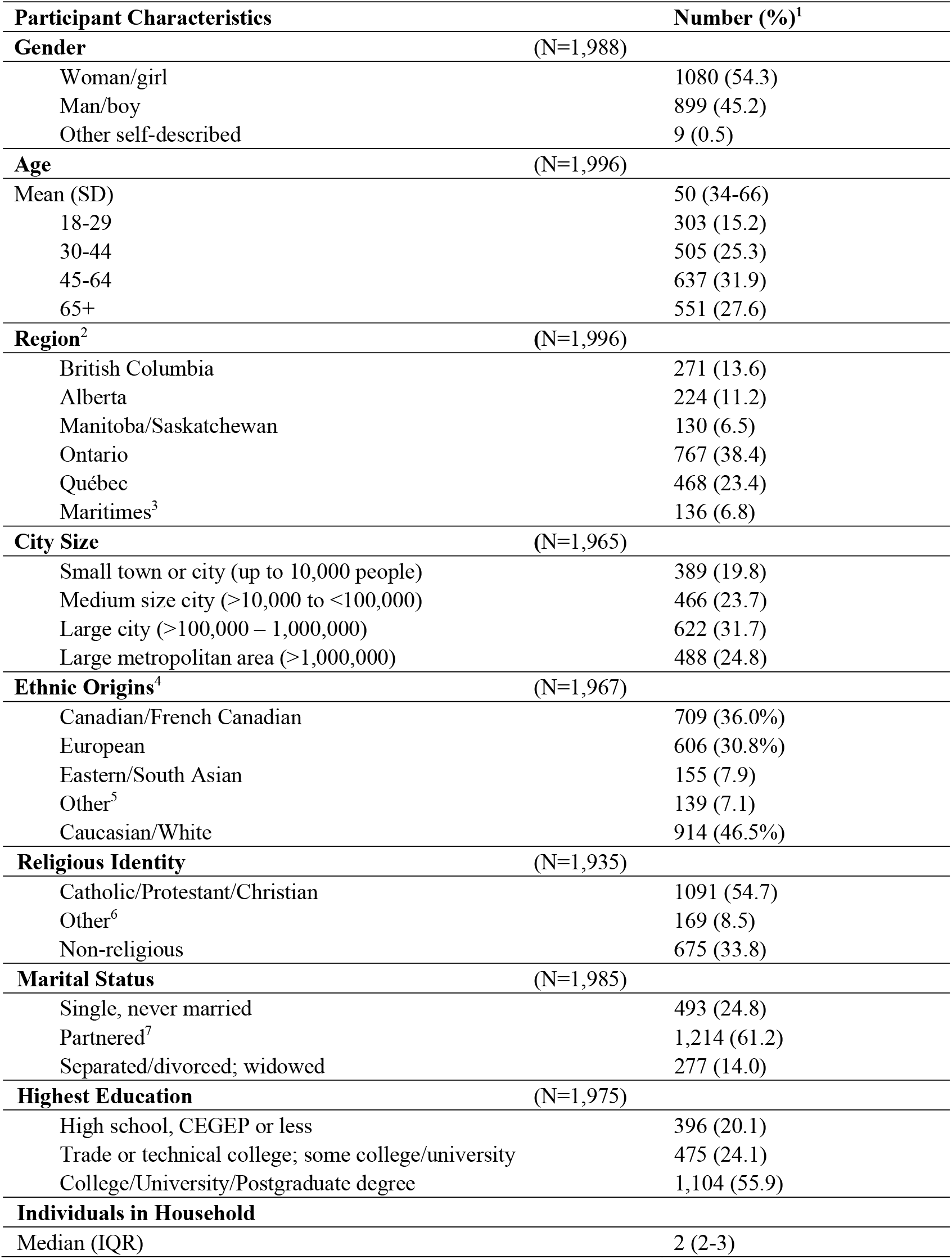
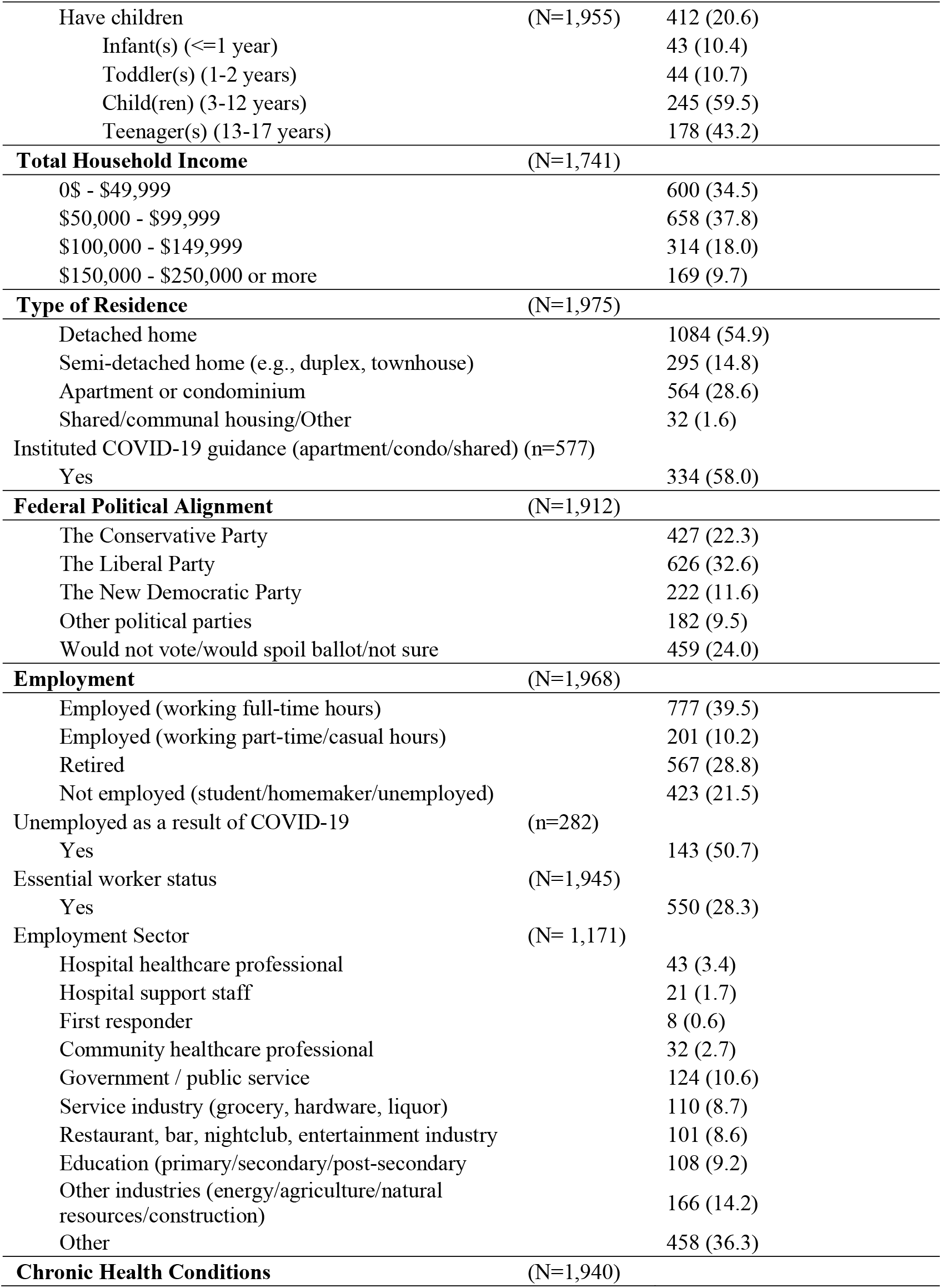
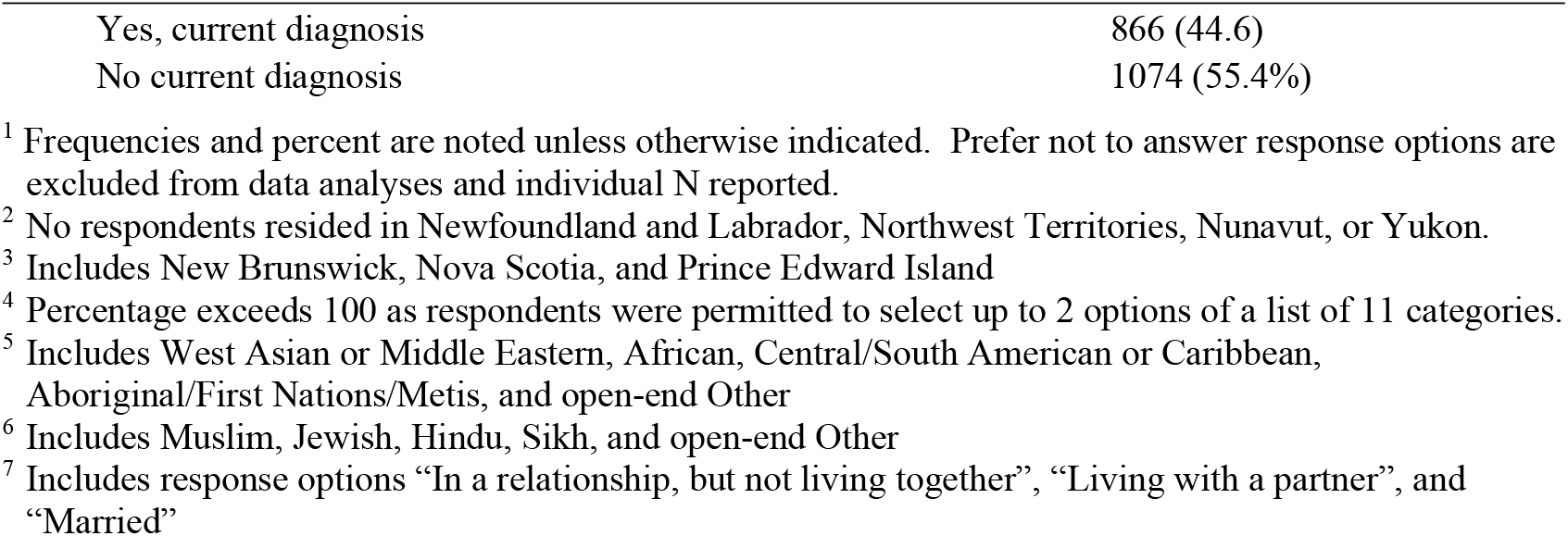
Respondent demographics (total sample size=1,996)

| **Participant Characteristics** | **Number (%)<sup>1</sup>** |
| --- | --- |
| **Gender** (N=1,988) |  |
| Woman/girl | 1080 (54.3) |
| Man/boy | 899 (45.2) |
| Other self-described | 9 (0.5) |
| **Age** (N=1,996) |  |
| Mean (SD) | 50 (34-66) |
| 18-29 | 303 (15.2) |
| 30-44 | 505 (25.3) |
| 45-64 | 637 (31.9) |
| 65+ | 551 (27.6) |
| **Region<sup>2</sup>** (N=1,996) |  |
| British Columbia | 271 (13.6) |
| Alberta | 224 (11.2) |
| Manitoba/Saskatchewan | 130 (6.5) |
| Ontario | 767 (38.4) |
| Québec | 468 (23.4) |
| Maritimes<sup>3</sup> | 136 (6.8) |
| **City Size** (N=1,965) |  |
| Small town or city (up to 10,000 people) | 389 (19.8) |
| Medium size city (>10,000 to <100,000) | 466 (23.7) |
| Large city (>100,000 – 1,000,000) | 622 (31.7) |
| Large metropolitan area (>1,000,000) | 488 (24.8) |
| **Ethnic Origins<sup>4</sup>** (N=1,967) |  |
| Canadian/French Canadian | 709 (36.0%) |
| European | 606 (30.8%) |
| Eastern/South Asian | 155 (7.9) |
| Other<sup>5</sup> | 139 (7.1) |
| Caucasian/White | 914 (46.5%) |
| **Religious Identity** (N=1,935) |  |
| Catholic/Protestant/Christian | 1091 (54.7) |
| Other<sup>6</sup> | 169 (8.5) |
| Non-religious | 675 (33.8) |
| **Marital Status** (N=1,985) |  |
| Single, never married | 493 (24.8) |
| Partnered<sup>7</sup> | 1,214 (61.2) |
| Separated/divorced; widowed | 277 (14.0) |
| **Highest Education** (N=1,975) |  |
| High school, CEGEP or less | 396 (20.1) |
| Trade or technical college; some college/university | 475 (24.1) |
| College/University/Postgraduate degree | 1,104 (55.9) |
| **Individuals in Household** |  |
| Median (IQR) | 2 (2-3) |

|  |  |  |
| --- | --- | --- |
| Have children | (N=1,955) | 412 (20.6) |
| Infant(s) (<=1 year) |  | 43 (10.4) |
| Toddler(s) (1-2 years) |  | 44 (10.7) |
| Child(ren) (3-12 years) |  | 245 (59.5) |
| Teenager(s) (13-17 years) |  | 178 (43.2) |
| **Total Household Income** | **(N=1,741)** |  |
| 0\$ - \$49,999 | | 600 (34.5) |
| \$50,000 - \$99,999 | | 658 (37.8) |
| \$100,000 - \$149,999 | | 314 (18.0) |
| \$150,000 - \$250,000 or more | | 169 (9.7) |
| **Type of Residence** | **(N=1,975)** |  |
| Detached home |  | 1084 (54.9) |
| Semi-detached home (e.g., duplex, townhouse) |  | 295 (14.8) |
| Apartment or condominium |  | 564 (28.6) |
| Shared/communal housing/Other |  | 32 (1.6) |
| Instituted COVID-19 guidance (apartment/condo/shared) (n=577) |  |  |
| Yes |  | 334 (58.0) |
| **Federal Political Alignment** | **(N=1,912)** |  |
| The Conservative Party |  | 427 (22.3) |
| The Liberal Party |  | 626 (32.6) |
| The New Democratic Party |  | 222 (11.6) |
| Other political parties |  | 182 (9.5) |
| Would not vote/would spoil ballot/not sure |  | 459 (24.0) |
| **Employment** | **(N=1,968)** |  |
| Employed (working full-time hours) |  | 777 (39.5) |
| Employed (working part-time/casual hours) |  | 201 (10.2) |
| Retired |  | 567 (28.8) |
| Not employed (student/homemaker/unemployed) |  | 423 (21.5) |
| Unemployed as a result of COVID-19 | (n=282) |  |
| Yes |  | 143 (50.7) |
| Essential worker status | (N=1,945) |  |
| Yes |  | 550 (28.3) |
| Employment Sector | (N= 1,171) |  |
| Hospital healthcare professional |  | 43 (3.4) |
| Hospital support staff |  | 21 (1.7) |
| First responder |  | 8 (0.6) |
| Community healthcare professional |  | 32 (2.7) |
| Government / public service |  | 124 (10.6) |
| Service industry (grocery, hardware, liquor) |  | 110 (8.7) |
| Restaurant, bar, nightclub, entertainment industry |  | 101 (8.6) |
| Education (primary/secondary/post-secondary) |  | 108 (9.2) |
| Other industries (energy/agriculture/natural resources/construction) |  | 166 (14.2) |
| Other |  | 458 (36.3) |
| **Chronic Health Conditions** | **(N=1,940)** |  |
| Yes, current diagnosis | 866 (44.6) |
| No current diagnosis | 1074 (55.4%) |

<sup>1</sup> Frequencies and percent are noted unless otherwise indicated. Prefer not to answer response options are excluded from data analyses and individual N reported.
<sup>2</sup> No respondents resided in Newfoundland and Labrador, Northwest Territories, Nunavut, or Yukon.
<sup>3</sup> Includes New Brunswick, Nova Scotia, and Prince Edward Island
<sup>4</sup> Percentage exceeds 100 as respondents were permitted to select up to 2 options of a list of 11 categories.
<sup>5</sup> Includes West Asian or Middle Eastern, African, Central/South American or Caribbean, Aboriginal/First Nations/Metis, and open-end Other
<sup>6</sup> Includes Muslim, Jewish, Hindu, Sikh, and open-end Other
<sup>7</sup> Includes response options “In a relationship, but not living together”, “Living with a partner”, and “Married”

### Public Perceptions

#### Severity

Most (n=1,236, 62.1%, 95%CI 59.9%-64.2%) respondents perceived COVID-19 to be a very serious problem in Canada though many rated it to be a less serious (n=914, 46.0%, 95%CI 43.7%-48.2%) problem than in other countries (Table A in S2 Appendix).

#### Concerns

More respondents were moderately or extremely concerned about a family member contracting COVID-19 (n=889, 45.3%, 95%CI 43.0%-47.5%) than were concerned about themselves contracting the disease (568/1,885 reported not believing they had contracted COVID-19, 30.1%, 95%CI 28.1%-32.2%) (Fig 1). In rating concerns about the impacts of COVID-19 on the health system, a greater propotion of respondents were moderately or extremely concerned that there would be insufficient personal protective equipment (PPE) for hospital staff to stay safe (n=1,024, 51.7%, 95%CI 49.5%-53.9%) compared to concerns about access to healthcare and availability of equipment to care for COVID-19 patients (Fig 1).

**Fig 1.**
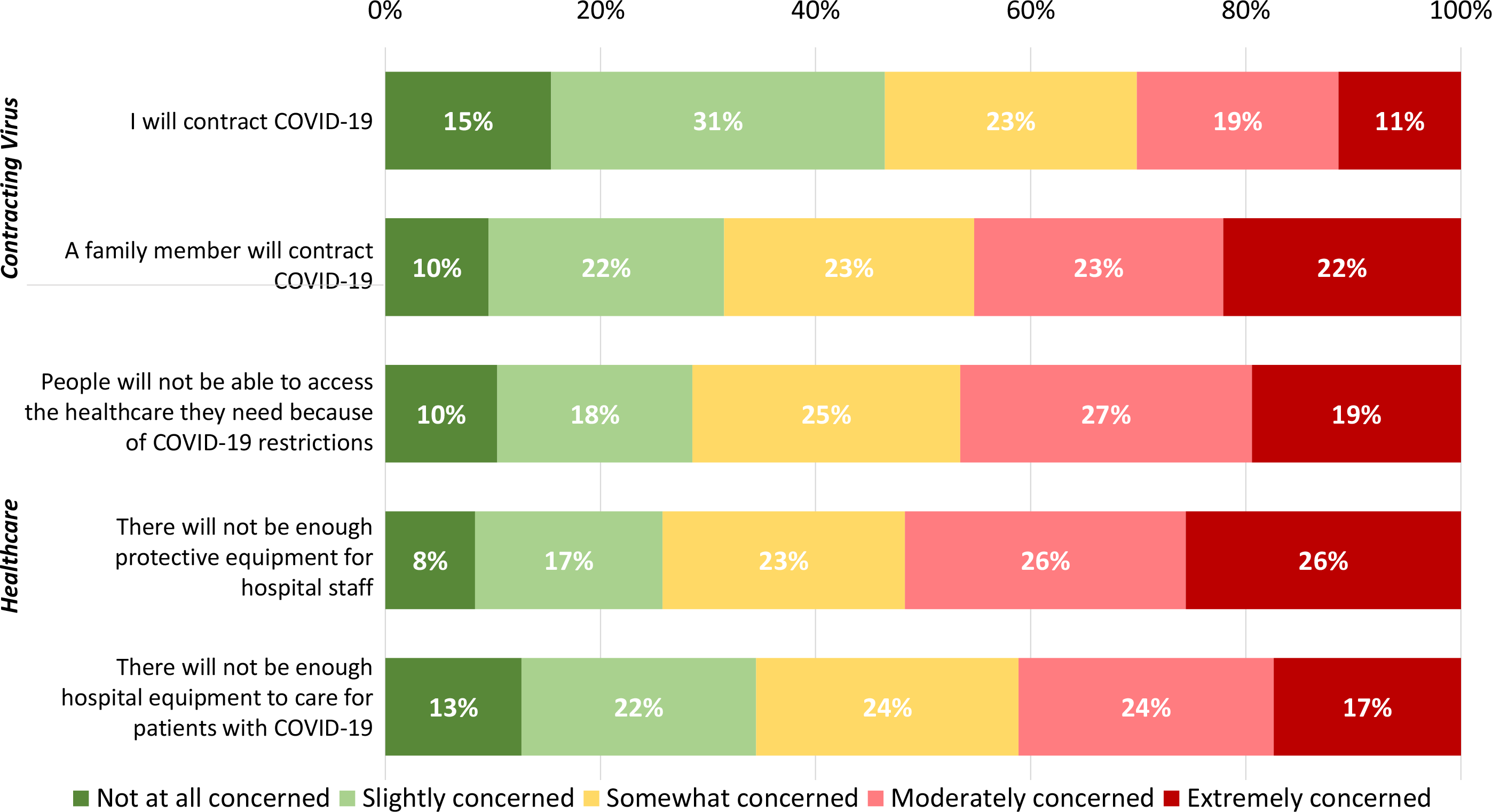
Respondents’ concerns about contracting the virus that causes COVID-19 and the impacts on healthcare.

#### Health

Just under half (n=898, 45.2%, 95%CI 43.0%-47.4%) of respondents agreed or strongly agreed that the pandemic was stressful; however, fewer (n=566, 28.5%, 95%CI 26.5%-30.5%) agreed or strongly agreed that it was something that made them feel helpless (Figure A in S2 Appendix). When asked to sequentially rate their past (start of 2020) and present health (physical, mental/emotional, social, economic, spiritual), respondents expressed experiencing declines in all dimensions of health with the largest decreases reported for social health (n= 964, 48.5%, 95%CI 46.3%-50.7%) and mental/emotional health (n=778, 39.1%, 95%CI 36.9%-41.2%)(Fig 2).

**Fig 2.**
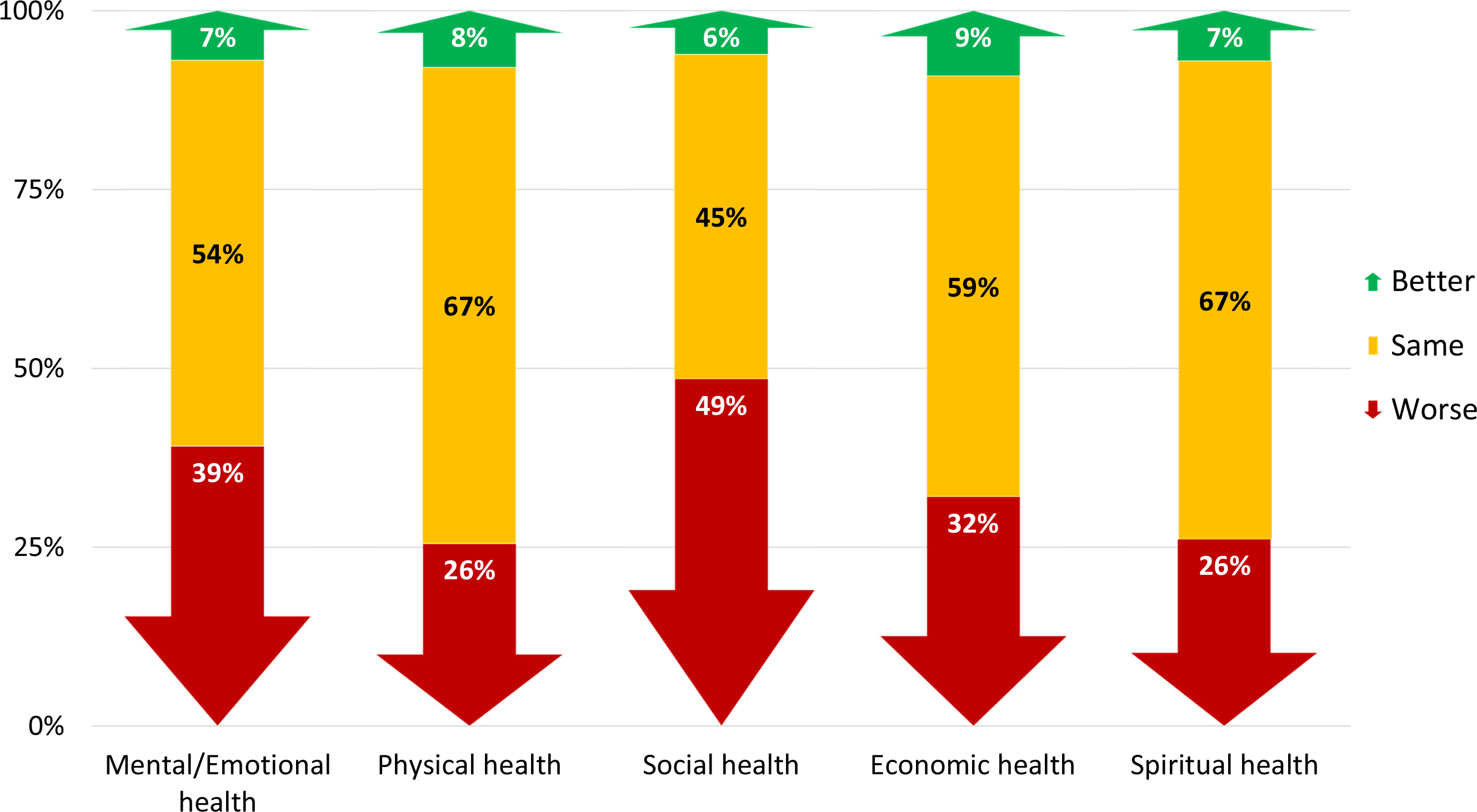
Difference in five domains of overall health at the start of 2020 compared to the time of questionnaire completion. Notes: Prefer not to answer responses are excluded from data analyses (range: n=5, 0.3% to n=107, 5.4%). Five-point scale ratings were poor, fair, good, very good, and excellent.

### Public Knowledge

#### Virus Transmission

The majority of respondents (n=1,741, 87.5%, 95%CI 86.1%-89.0%) rated their understanding of how the virus was spread as good (n=629, 31.6%, 95%CI 29.5%-33.7%), very good (n=793, 39.9%, 95%CI 37.7%-42.0%), or excellent (n=319, 16.1%, 95%CI 14.4%-17.7%). Fig 3 shows respondents’ level of agreement to a series of statements about the transmission of the virus that causes COVID-19. The highest consensus among respondents was in agreeing or strongly agreeing that people can be infected with COVID-19 and not show any symptoms (n=1,713, 86.5%, 95%CI 84.9%-88.0%). There was greater variability across respondents in their degree of agreement to other knowledge-based statements (Fig 3).

**Fig 3.**
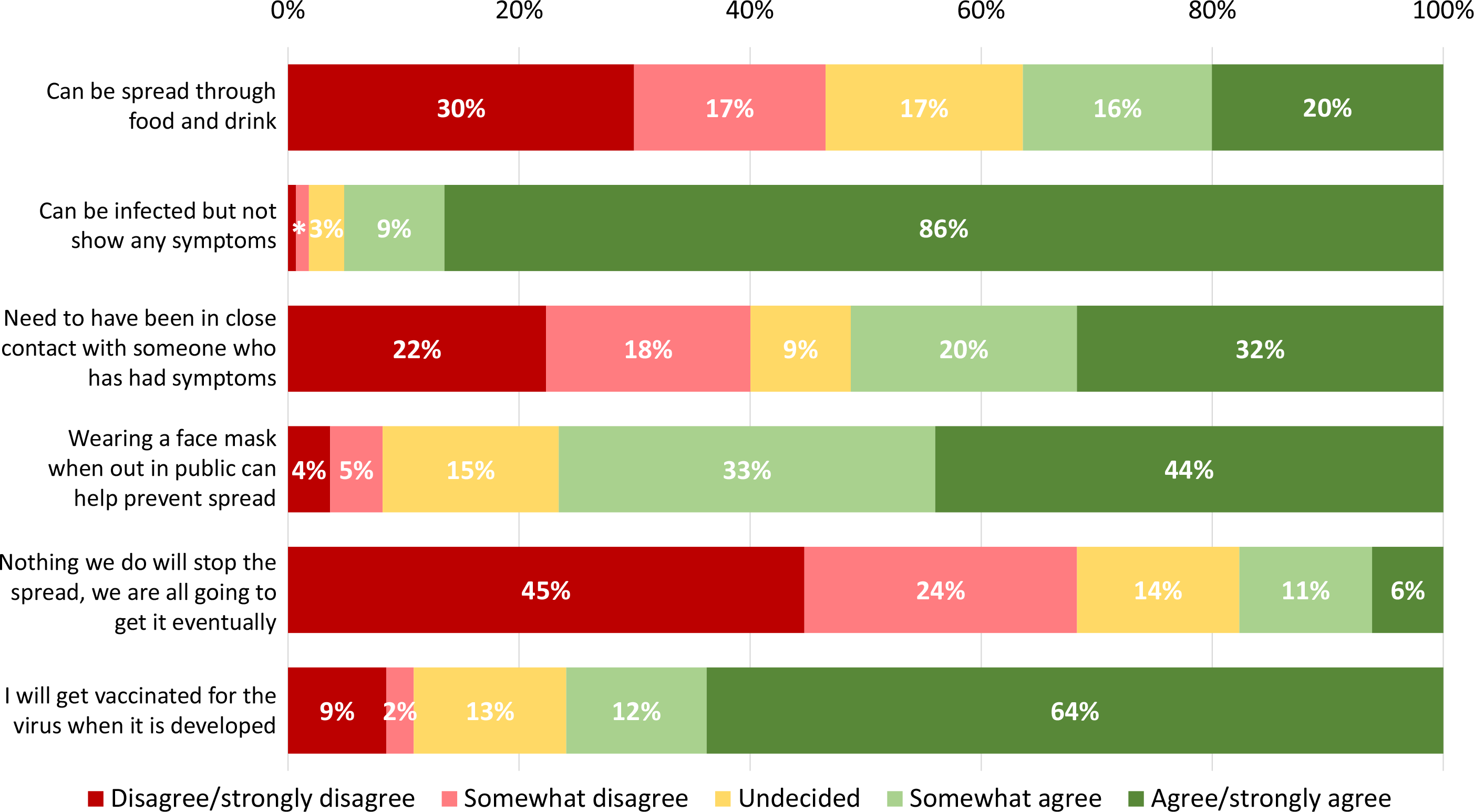
Respondents’ understanding of virus transmission and mitigation. Notes: Prefer not to answer response options are excluded from data analyses (range: n=15, 0.8% to n=146, 7.3%). * Percentage for somewhat disagree = 1%; percentage for disagree/strongly disagree = 1%.

#### Sources of Information

Over half (n=1,345, 67.9%, 95%CI 65.8%-69.9%) of respondents reported searching for information about COVID-19 once per day or more and predominantly accessing and trusting Canadian over American or other international sources for information. The top accessed source was Canadian news-based television, print, or websites (n=1,488, 75.6%, 95% CI 73.6%-77.5%) (Fig 4). The lowest rated sources for COVID-19 information included social media posts from influencers or celebrities (n=1,039, 54.8% selected as least trusted, 95% CI 52.5%-57.1%) and American news television, print, and websites (n=711, 50.4% selected as source of misinformation, 95% CI 47.7%-53.0%). Consistent with valuing Canadian sources, respondents most frequently reported going directly to government or health authority sources (n=979, 50.6%, 95% CI 48.4%-52.8%) to verify information (Figure B in S2 Appendix).

**Fig 4.**
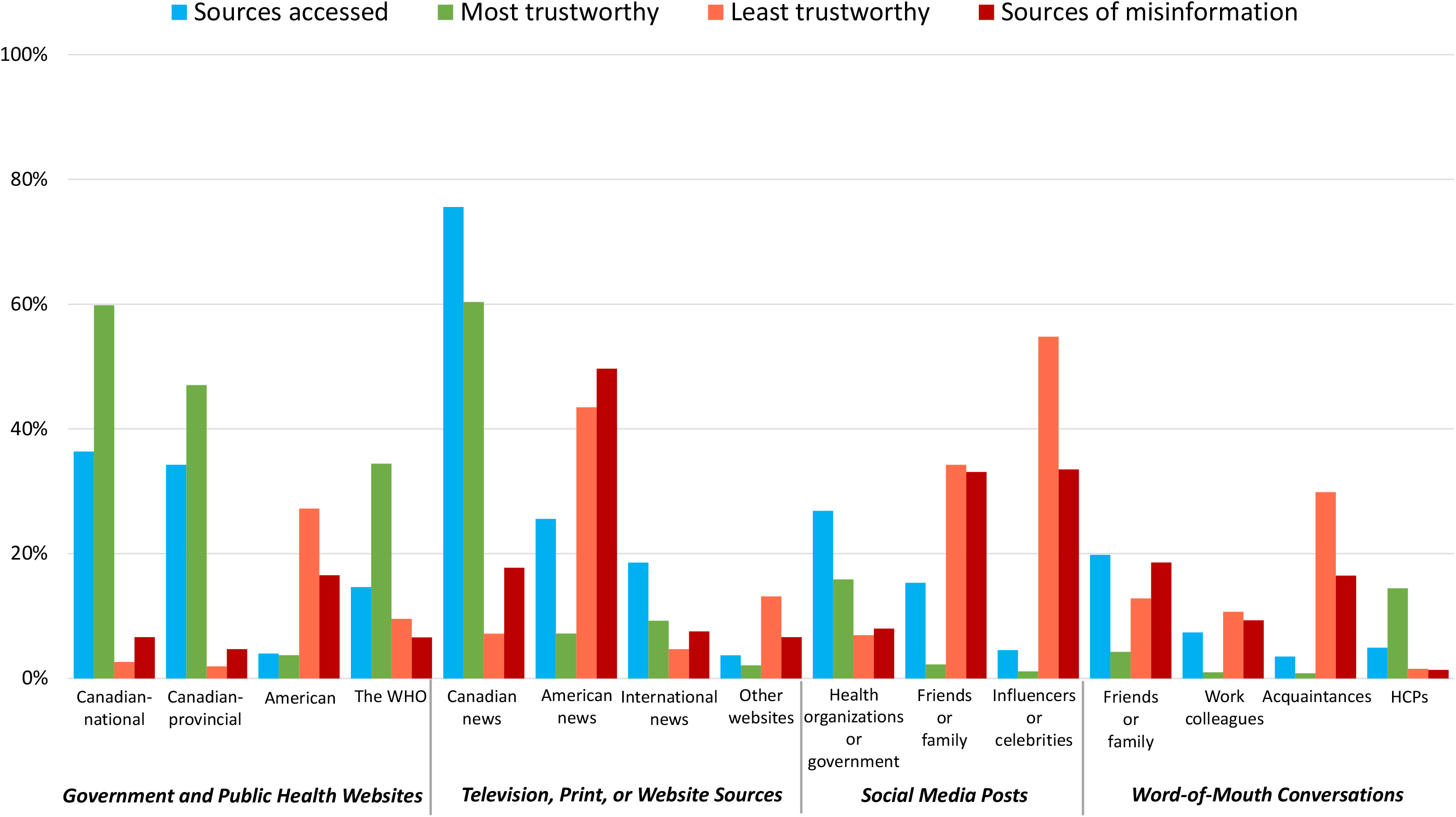
Information sources accessed, selected as most trustworthy, least trustworthy, and sources of misinformation indicated by respondents. Notes: Prefer not to answer responses are excluded from analysis (range: n=5, 0.3% to n=99, 5.0%). Canadian news is a combined category of Canadian television news, Canadian newspapers/magazines, and Canadian news websites American news is a combined category of American television news, American newspapers/magazines, and American news websites HCPs=healthcare providers; WHO = World Health Organization

Half of respondents surveyed (n=1,017, 51.3%, 95% CI 49.1%-53.5%) agreed or strongly agreed that they were able to find the kind of information they want about COVID-19 (Figure C in S2 Appendix). Information about COVID-19 infection rates dominated respondent’s searches (n=1,414, 71.5%, 95% CI 69.5%-73.5%) (Figure D in S2 Appendix), while information about vaccines and treatments were most frequently (n=933, 48.9%, 95% CI 46.7%-51.2%) cited as topics of misinformation (Figure D in S2 Appendix) from those who reported having seen or heard incorrect or misleading information related to COVID-19 during the previous two weeks (n=1,520, 75.3%, 95% CI 73.4%-77.3%). Yet, only half (n=937, 47.4%, 95% CI 45.2%-49.6%) of respondents felt moderately or extremely confident that they could identify incorrect or misleading information about COVID-19 (Figure E in S2 Appendix), and comparable numbers reported being uncertain (n=455, 23.0%, 95% CI 21.2%-24.9%) or agreeing (n=634, 32.1%, 95% CI 30.0%-34.2%) that they find it hard to determine if an information source was trustworthy or not (Figure E in S2 Appendix).

### Public Behaviors

#### Mitigation and Containment

Just under half of respondents indicated they were in self-isolation (n=842, 43.4%, 95% CI 41.2%-45.6%). Of those who were not self-isolating (n=1,144), the vast majority (n=1,083, 95.1%, 95% CI 93.8%-96.4%) reported that they practiced physical distancing always (n=783, 68.8%, (95% CI 66.0%-71.5%) or often (n=300, 26.3%, 95% CI 23.7%-28.9%). Furthermore, many (n=814, 41.0%, 95% CI 38.9%-43.2%) respondents felt that they could reasonably sustain their *current level* of physical distancing longer than six months (or as long as needed) (Fig 5). Self-reported distancing behaviors were consistent with respondent perceptions of ‘self’ as effective agents to prevent the spread of the virus, with most (n=1,380, 69.7%, 95% CI 67.7%-71.8%) agreeing or strongly agreeing that they were doing a good job at preventing the spread of the virus with changes to their behavior; about one-third (n=677, 34.9%, 95% CI 32.7%-37.0%) agreed or strongly agreed that they were doing a better job than other people (Figure E in S2 Appendix). Respondents (mean age of 50) most commonly perceived teenagers as least consistently practicing physical distancing (n=855, 43.2%, 95% CI 41.0%-45.4%) while identifying middle-aged adults (n=786, 39.6%, 95%CI 37.4%-41.8%) and seniors (n=744, 37.5%, 95%CI 35.4%-39.6%) as most consistently practicing physical distancing (Figure F in Appendix S2).

**Fig 5.**
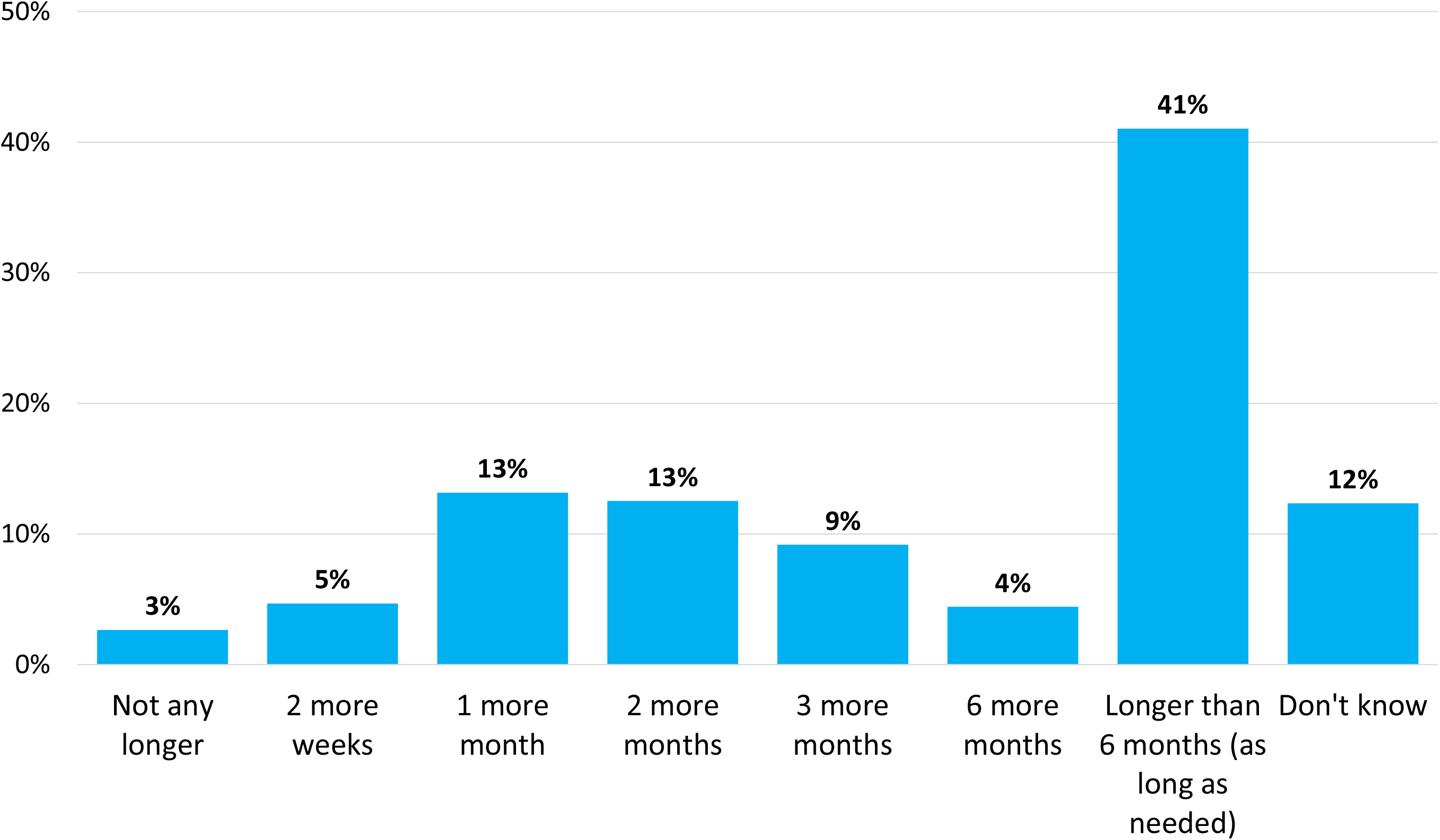
Proportion of respondents who indicated how long they believe they could sustain their current level of physical distancing. Note: Prefer not to answer response options are excluded from data analyses (n=12, 0.6%).

#### Motivations

The most frequently selected motivations (Fig 6) for those who reported self-isolating or physical distancing were to protect oneself (n=1,602, 81.0%, 95% CI 79.2%-82.7%), to protect other people in one’s household (n=970, 49.1%, 95% CI 46.8%-51.3%) and to protect other members of the general public (n=962, 48.6%, 95% CI 46.4%-50.8%). Three-quarters (n=1,436, 75.8%, 95% CI 73.9%-77.8%) of respondents reported that they would get vaccinated for the virus when a vaccine became available.

**Fig 6.**
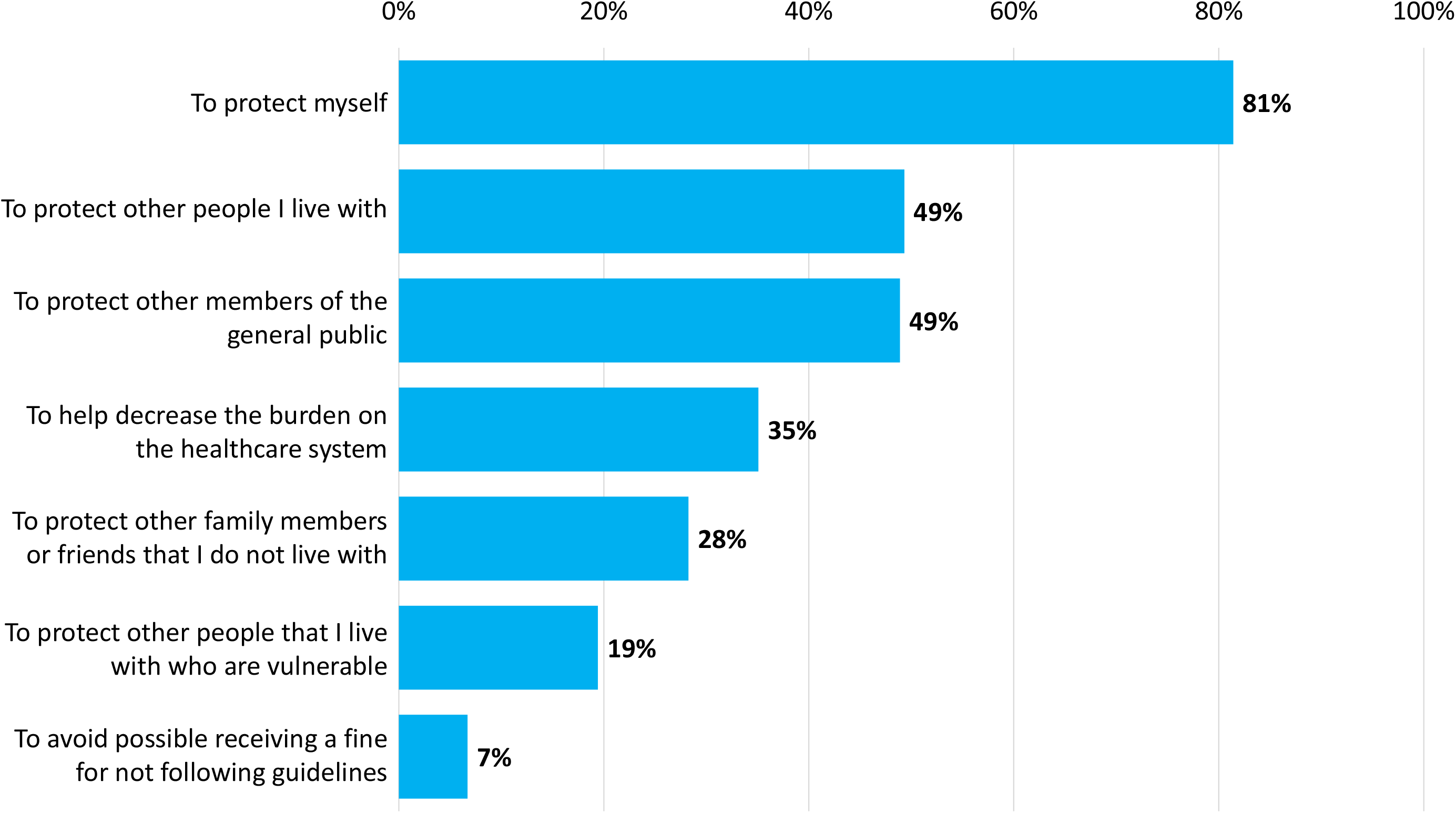
Respondents’ motivations for practicing either self-isolation (n=852) or physical distancing (n=1,126). Notes: Prefer not to answer response options are excluded from data analyses (n=6, 0.7% and n=4, 0.4%, respectively).

#### Regional Differences

Sub-group analyses revealed several regional differences (see S1 Table) mostly with differences between Ontario and Québec compared to other provinces. A greater proportion of respondents in Ontario rated COVID-19 as a very serious problem (p<0.001) and expressed higher rates of concern about the impact of COVID-19 on hospitals (e.g. lack of PPE, p<0.001) and patients (e.g. limited access to necessary services, p<0.001) than respondents in other provinces. Respondents from Ontario and Québec were more concerned than those in other regions about contracting COVID-19 (p=0.002) and about a family member contracting COVID-19 (p=0.021); those in Québec were more likely to have close friends who tested positive for COVID-19 (p=0.011). A greater proportion of respondents in Ontario and the Maritime provinces agreed or strongly agreed that the pandemic is stressful (p<0.001), while a greater proportion of Québec residents felt helpless (p<0.001). There were no regional differences in perceptions of physical, mental/emotional, social, or economic health. Respondents in Québec felt least knowledgeable about how the virus is spread (p<0.001). In addition, they tended to access Canadian news more often than those in other regions, while those in BC and Ontario accessed American and international sources more often than others (p<0.001). Significantly fewer residents (52%) in Québec agreed or strongly agreed that they would get vaccinated compared to participants from other regions (range 64%-73%) (p<0.001).

## Discussion

The COVID-19 pandemic has substantially altered many aspects of public life, yet little is known about the perspectives and experiences of broader populations. Our study provides a national cross-sectional description of public perceptions, knowledge and behaviors related to COVID-19 in the context of the evolving pandemic, adding to survey data published early in the outbreak.(16-18). Our data suggest that Canadians are concerned about the threat of COVID-19 to the healthcare system, to themselves and their family members, and that they consider the ongoing pandemic a serious problem on both national and international levels. The main findings of the survey include the negative impact of the pandemic on Canadians’ perceptions of their health, the frequent searching for up-to-date information about COVID-19 (largely via Canadian based sources), and current and future perceived desire and ability of the public to comply with public health recommendations (e.g. physical distancing, vaccination for COVID-19 if/when available). To our knowledge, this is the first national survey in Canada to comprehensively assess multiple domains (perceptions, knowledge, and behaviors) important to understanding the public’s response to the ongoing pandemic.

### Health and Well-being

We found that overall health has been markedly impacted by pandemic conditions, and that this is irrespective of personal infection with COVID-19. In fact, very few of our respondents reported testing positive for COVID-19, yet many perceived that aspects of their overall health had deteriorated, particularly mental/emotional and social health. This is further evidenced in high agreement among our respondents that the pandemic is stressful. The need to assess and respond to health impacts beyond infection with SARS-Cov-2 has been increasingly recognized as a critical part of pandemic response.(31-37) For example, dramatic shifts in routines, livelihoods and behaviors during quarantine, coupled with the unfulfilled basic need for human connection,(33) have been described as significant threats to mental health and well-being. In addition, findings from surveys commissioned by the UK Academy of Medical Sciences (AMS) and the charity MQ: Transforming Mental Health through Research reported widespread public concern about isolation, loneliness, practical aspects of life (e.g. finances), and general negative feelings, and provided groundwork for the collaborative development of sweeping research priorities to improve these conditions.(34) In our survey, fewer respondents reported that the pandemic makes them feel helpless, suggesting some resiliency to the detrimental circumstances the pandemic has produced.

### Information, Misinformation and Effective Messaging

The media’s role in disseminating information that will concurrently educate and motivate public behaviors in accordance with recommended guidance and avoid creating undue stress, skepticism, or rebuff of guidelines is a critical factor in navigating pandemic response.(32, 34, 38) Our study found that the public frequently searches for information about COVID-19 and is primarily getting information from domestic news sources, including television, print, and websites that are not government or public health agency websites. Respondents in our study also view news sources as equally credible to national government and public health websites. This finding suggests that public health officials should view mainstream media, and in particular television, as important promoters or messengers of COVID-19-related information. Given this, it is crucial for mainstream media to take this responsibility seriously to ensure accurate information is conveyed. At the same time, perceptions of trust may be moderated by other factors not accounted for in this survey, such as perceived congruence between government guidelines and impact reducing virus spread. In our survey, respondents from Ontario and Québec reported the least amount of trust in Canadian government and news sources and these were also the same provinces with the highest number of confirmed COVID-19 cases in Canada (32% and 51%, respectively).

Much attention has been paid to the proliferation of information about COVID-19, raising concerns about parallel increases in misinformation. We found that a substantial proportion of respondents value science-based sources (e.g. government websites) which may explain high rates of self-reported behavior change to prevent virus spread. This correlates with other public opinion data;(39) however, about half of our respondents still expressed only moderate levels of confidence in being able to identify misleading information or determine if an information source is trustworthy (Figure D in S2 Appendix). Of note, many respondents indicated that they do not view American news sources as trustworthy, and more specifically, see it as a source of misinformation. Familiarity with and interest in context-specific information may influence respondents’ perceptions of credibility. Social media posts were also commonly identified as untrustworthy, however, these perceptions ranged depending on who was sharing the information. Posts from family and friends or influencers were viewed as less trustworthy than posts from government or public health agencies. As a quick-response platform with open posting and limited moderation, misinformation is easily spread on social media.(39-41) While some social media platforms (e.g. Facebook, Twitter and Instagram) have increased efforts to monitor and remove incorrect or harmful information related to COVID-19 in an attempt to reduce public consumption of misinformation, the effectiveness of these efforts is currently unknown.(42) Moreover, efforts to better understand how individuals may proactively limit their exposure to misinformation, identify misinformation, and fact check information are needed. We found that our respondents most frequently fact checked their information using government and public health websites (51%) and scientific articles (30%).

### Compliance with Public Health Recommendations

The vast majority of respondents in our study reported practicing a high level of physical distancing, and a surprisingly high number felt that they could maintain this for a long period of time (6 months or more) if necessary. This finding is somewhat unexpected given the high level of reported self-isolation amongst our respondents. Although it may be that not all respondents clearly understood the difference between self-isolation and physical distancing, it is evident that most were motivated to limit social and physical interactions as a means to protect themselves and others from becoming infected with COVID-19. The lower than predicted infection rates in many countries has been credited largely to the high public compliance of mandated preventative measures. However, this comes at a price, including significant global economic losses.(43) In our survey, 14% of respondents reported unemployment as a result of the pandemic, and 34% of all respondents reported worse economic health.

In contrast to respondents’ positive association to physical distancing recommendations, we report slightly lower numbers of respondents who intend to receive a COVID-19 vaccine once available as compared to other recent surveys.(44) While this is another somewhat unanticipated finding given the reported propensity of respondents to access and trust sources considered ‘reputable’ (e.g., public health agencies), individual and social determinants of vaccination are wide-ranging.(45-48) Previous research has highlighted that the media can both hinder(49, 50) and enhance(50) vaccination uptake. To optimize potential future vaccine uptake, public health agencies should align key messaging with public perceptions, concerns, and information needs (e.g. preferred sources),(51, 52) tailoring by jurisdiction. For example, in our study, respondents from the province with the highest number of COVID-19 cases (Québec) were significantly less likely to report that they plan to get vaccinated (S1 Table) and reported the least amount of trust in Canadian government and news sources. Such complexities must be taken seriously if we are to ensure that public health recommendations are effectively communicated.

### Limitations

Our survey has limitations. Although providing a broad snapshot of population, cross-sectional surveys capture relevant data at a single moment in time. In a rapidly changing landscape, it is expected that self-reported perceptions and behaviors would change with new information. The use of serial surveys(13, 14) is one strategy to strengthen cross-sectional survey designs. At the same time, our study provides useful descriptive data at the height of the pandemic in Canada. Subsequent qualitative methodologies will further enrich our understanding of public actions and reactions to the COVID-19 pandemic. Second, as we elected to set a survey response quota, we are not able to determine a response rate. While there is a risk of non-response bias, the rapid collection of responses to reach our 2,000 quota (five days) and methodological strengths in our design (rigorous development including pre-testing and device agnosticism, large sample size, population representation and weighting by age, sex, and region) outweigh this limitation. Third, differences in public perceptions, knowledge, and behaviors that may be associated with socio-demographic factors such as age and gender were not addressed in this manuscript but will be the focus of future investigation. Finally, though overall results may be affected by larger numbers of respondents from Canada’s two largest provinces (Ontario and Québec), the weighting ensures results accurately reflect the actual regional populations within Canada. At the same time, regional differences should be cautiously interpreted as we did not adjust for multiple comparisons.

## Conclusions

We conducted a national survey including a representative sample of the Canadian public to assess overall perceptions, knowledge, and behaviors related to the COVID-19 pandemic. Our results highlight the impact of the pandemic on individual perceptions of health which may be further exacerbated by salient concerns around risks of infection, healthcare safety, and access. We found that knowledge about COVID-19 is largely acquired through domestic news sources, which may explain high self-reported compliance with prevention measures. The findings of this study should be used to inform public health communications during COVID-19 and future pandemics.

## Data Availability

All relevant data are within the paper and its Supporting Information files.

## Funding

This work was supported by the Canadian Institutes of Health Research Canadian 2019 Novel Coronavirus (COVID-2019) Rapid Research Funding Opportunity – Operating Grant (grant number RN420046-439965) to JPL. The funders had no role in study design, data collection and analysis, decision to publish, or preparation of the manuscript.

## Declaration of Interests

JPL is primary investigator, KF, AS, SBA, KEAB, AFR, SK, SL, SM, DJN, BR, HTS are co-investigators, on the grant from the Canadian Institutes of Health Research (CIHR) for the submitted work; AFR has received research grants from the Canadian Institutes of Health Research (CIHR), the Natural Sciences and Engineering Research Council (NSERC), and Hamilton Academic Health Sciences Organization and is President of the Canadian Sepsis Foundation, HTS is supported by an Embedded Clinician Re-searcher Award from the CIHR; all authors declare no other relationships or activities that could appear to have influenced the submitted work.

## Author Contributions

Funding Acquisition – JPL

Conceptualization – JPL, K, HTS

Data curation and analysis – AS

Investigation – JPL, RBM, KP, AS, LWB

Methodology – JPL, KF, RBM, KP, AS, LWB, SK, DJN, HTS

Project Administration/Supervision – JPL, KF, RBM, LWB

Writing Original Draft / Visualization – JPL, KF, RBM, KP, AS, EES, HTS

Writing Review and Editing – JPL, KF, RBM, KP, AS, EES, LWB, SBA, KEAB, AFR, SK, SL, SM, DJN, BR, HTS

All authors critically revised the manuscript for important intellectual content. All of the authors read and approved the final version to be published and agreed to be accountable for all aspects of the work.

## Supporting Information

S1 Appendix. Socio-Cultural Implications of COVID-19, Public Perceptions Survey.

S1 Fig. Survey content domains and sub-domains.

S2 Appendix. Additional data figures and tables

Table A. Self-reported perceptions, knowledge, and behaviors.
Figure A. Perceived psychological impact of COVID-19 and sufficiency of government response.
Figure B. Proportion of respondents who used various strategies to fact check misinformation
Figure C. Respondents’ evaluation of information seeking and preventative behaviors.
Figure D. Topics searched for and topics identified as misinformation.
Figure E. Confidence in self and others ability to identify misinformation.
Figure F. Perceptions of age groups most and least consistently practicing physical distancing.

S1 Table. Regional differences in perceptions, knowledge, and behaviors related to COVID-19.

## Notes

### Competing Interest Statement

JPL is the primary investigator, KF, AS, SBA, KEAB, AFR, SK, SL, SM, DJN, BR, and HTS are co-investigators on a grant from the Canadian Institutes of Health Research (CIHR) for the submitted work; AFR has received research grants from the Canadian Institutes of Health Research (CIHR), the Natural Sciences and Engineering Research Council (NSERC), and Hamilton Academic Health Sciences Organization and is President of the Canadian Sepsis Foundation; HTS is supported by an Embedded Clinician Researcher Award from the CIHR; all authors declare no other relationships or activities that could appear to have influenced the submitted work.

### Author Declarations

Dalhousie University (#2020-5121) and the University of Calgary (#20-0538) Research Ethics Boards approved this study.

